# Comparison of outcomes from COVID infection in pediatric and adult patients before and after the emergence of Omicron

**DOI:** 10.1101/2021.12.30.21268495

**Authors:** Lindsey Wang, Nathan A. Berger, David C. Kaelber, Pamela B. Davis, Nora D. Volkow, Rong Xu

**Affiliations:** Center for Artificial Intelligence in Drug Discovery, Case Western Reserve University School of Medicine, Cleveland, OH, USA; Center for Science, Health, and Society, Case Western Reserve University School of Medicine, Cleveland, OH, USA; Case Comprehensive Cancer Center, School of Medicine, Case Western Reserve University, Cleveland, OH, USA; The Center for Clinical Informatics Research and Education, The MetroHealth System, Cleveland, OH, USA; Center for Community Health Integration, School of Medicine, Case Western Reserve University, Cleveland, OH, USA; National Institute on Drug Abuse, National Institutes of Health, Bethesda, MD, USA

## Abstract

**Background:** The Omicron SARS-CoV-2 variant is rapidly spreading in the US since December 2021 and is more contagious than earlier variants. Currently, data on the severity of the disease caused by the Omicron variant compared with the Delta variant is limited. Here we compared 3-day risks of emergency department (ED) visit, hospitalization, intensive care unit (ICU) admission, and mechanical ventilation in patients who were first infected during a time period when the Omicron variant was emerging to those in patients who were first infected when the Delta variant was predominant.

**Method:** This is a retrospective cohort study of electronic health record (EHR) data of 577,938 first-time SARS-CoV-2 infected patients from a multicenter, nationwide database in the US during 9/1/2021–12/24/2021, including 14,054 who had their first infection during the 12/15/2021–12/24/2021 period when the Omicron variant emerged (“Emergent Omicron cohort”) and 563,884 who had their first infection during the 9/1/2021–12/15/2021 period when the Delta variant was predominant (“Delta cohort”). After propensity-score matching the cohorts, the 3-day risks of four outcomes (ED visit, hospitalization, ICU admission, and mechanical ventilation) were compared. Risk ratios, and 95% confidence intervals (CI) were calculated.

**Results:** Of 14,054 patients in the Emergent Omicron cohort (average age, 36.4 ± 24.3 years), 27.7% were pediatric patients (<18 years old), 55.4% female, 1.8% Asian, 17.1% Black, 4.8% Hispanic, and 57.3% White. The Emergent Omicron cohort differed significantly from the Delta cohort in demographics, comorbidities, and socio-economic determinants of health. After propensity-score matching for demographics, socio-economic determinants of health, comorbidities, medications and vaccination status, the 3-day risks in the Emergent Omicron cohort outcomes were consistently less than half those in the Delta cohort: ED visit: 4.55% vs. 15.22% (risk ratio or RR: 0.30, 95% CI: 0.28-0.33); hospitalization: 1.75% vs. 3.95% (RR: 0.44, 95% CI: 0.38-0.52]); ICU admission: 0.26% vs. 0.78% (RR: 0.33, 95% CI:0.23-0.48); mechanical ventilation: 0.07% vs. 0.43% (RR: 0.16, 95% CI: 0.08-0.32). In children under 5 years old, the overall risks of ED visits and hospitalization in the Emergent Omicron cohort were 3.89% and 0.96% respectively, significantly lower than 21.01% and 2.65% in the matched Delta cohort (RR for ED visit: 0.19, 95% CI: 0.14-0.25; RR for hospitalization: 0.36, 95% CI: 0.19-0.68). Similar trends were observed for other pediatric age groups (5-11, 12-17 years), adults (18-64 years) and older adults (≥ 65 years).

**Conclusions:** First time SARS-CoV-2 infections occurring at a time when the Omicron variant was rapidly spreading were associated with significantly less severe outcomes than first-time infections when the Delta variant predominated.

## Introduction

Omicron SARS-CoV-2 variant is rapidly spreading in the United States (US) and its prevalence increased from 22.5% between 12/12/2021–12/18/2021 to 58.6% between 12/19/2021– 12/25/2021, according to CDC’s national SARS-CoV-2 genomic surveillance program^1^. The Omicron variant is more contagious than the Delta variant^2^. However, the data on the severity of the disease caused by the Omicron variant is scarce and incomplete^3^. Reports from South Africa^4^, Scotland^5^, and England^6^ showed lower rates of hospitalization following Omicron infection compared with infections from the Delta variant. Currently, data on the broader spectrum of disease severity caused by the Omicron variant compared with the Delta variant in the US is lacking. Here we compared the risks of ED visit, hospitalization, ICU admission, and mechanical ventilation in patients who were first infected during the period when the Omicron variant was rapidly spreading in the US (“Emergent Omicron cohort”) and compared them to those in similar patients who were first infected during the period when the Delta variant predominated (“Delta cohort”) through a retrospective study of a large, geographically diverse database of patient electronic health record (EHR) data in the US. Outcomes in pediatric patients (age 0-4, 5-11 and 12-17 years), adults (18-64 years) and older adults (≥ 65 years) were examined.

## METHODS

### Study population

This study used the TriNetX Analytics network platform that contains de-identified HER data of 84.5 million unique patients from 63 health care organizations of both inpatient and outpatient settings across the US^7^. TriNetX Analytics provides web-based secure access to patient EHR data from hospitals, primary care, and specialty treatment providers, covering diverse geographic locations, age groups, racial and ethnic groups, income levels, and insurance types. Although the data are fully de-identified, end-users can use built-in statistical functions to perform patient-level data analysis, including cohort selection, propensity-score matching, time trend analysis, outcome research, among others. Because this study only queried statistics of de-identified patient records through web-applications and did not involve retrieval, storage, collection, use, or transmittal of individually identifiable data, Institutional Review Board approval and informed consent was not needed or sought.

The study population comprised three cohorts of patients with SARS-CoV-2 infections: (a) the Emergent Omicron cohort (n = 14,054) – patients who had their first SARS-CoV-2 infections between 12/15/2021–12/24/2021, based on documented evidence in the EHR and had not experienced previous SARS-CoV-2 infection. The CDC’s national genomic surveillance program reports that the prevalence of the Omicron variant in the US increased from 22.5% between 12/12/2021–12/18/2021 to 58.6% between 12/19/2021–12/25/2021^1^; (b) the Delta cohort (n = 563,884) – patients who had their first SARS-CoV-2 infections between 9/1/2021– 11/15/2021 based on documented evidence in the EHR and had not experienced previous SARS-CoV-2 infection. Based on CDC data, Delta was the predominant variant at that time (98.6-99.1%)^1^; (c) the Delta-2 cohort (n = 77,692) – patients who had their first SARS-CoV-2 infections between 11/16/2021–11/30/2021, immediately before the Omicron variant was detected in the US, and had no prior SARS-CoV-2 infection. Delta was the predominant variant at this time (98.4-99.0%)^1^. This second Delta cohort was created to control for later time periods and shorter window of infection.

The status of SARS-CoV-2 infection was based on the ICD-10 diagnosis code of “COVID-19” (U07.1) or lab-test confirmed presence of “SARS coronavirus 2 and related RNA” (9088). The status of outcomes was based on the Current Procedural Terminology (CPT) relevant codes for ED visit (“Emergency Department Visits”, code 1013711), hospitalization (“hospital inpatient services”, code: 013659), ICU admission (“Critical Care Services”, code: 1013729), and mechanical ventilation (“Respiratory ventilation”, codes: 5A1935Z, 5A1945Z, 5A1955Z, 5A09357, 5A09457, 5A09557).

### Statistical analysis

We tested whether the Emergent Omicron cohort had severity in outcomes different from the Delta cohort within the three days following infection. The two cohorts were propensity-score matched (1:1 using a nearest neighbor greedy matching with a caliper of 0.25 times the standard deviation) for demographics (age, gender, race/ethnicity); adverse socioeconomic determinants of health (assessed by ICD-10 codes “Z55-Z65” for “Persons with potential health hazards related to socioeconomic and psychosocial circumstances”) that include employment, housing, education, and economic circumstances; transplant procedures; comorbidities relevant to COVID-19 risks or outcomes^8,9^ including hypertension, heart diseases, cerebrovascular diseases, cancer, obesity, type 2 diabetes, chronic respiratory diseases, chronic kidney diseases, liver diseases, HIV infection, dementia, substance use disorders, depression and anxiety (assessed by ICD-10 codes); behavioral factors (tobacco smoking, alcohol drinking) (assessed by one or more encounter based on ICD-10 codes); COVID-19-related medications^10^ including remdesivir, dexamethasone, hydrocortisone, tocilizumab, fluvoxamine, and fluoxetine (assessed by RxNorm codes); and vaccination status documented in patient EHRs (Pfizer, Moderna, J&J) and booster (3^rd^ or booster shorts of Pfizer or Moderna mRNA vaccines) (assessed by CPT codes).

Four outcomes (ED visit, hospitalization, ICU admission, mechanical ventilation) were examined in the time window from the first day of SARS-CoV-2 infection identification to 3 days after infection, with all analyses being performed on 12/27/2021. We selected the 3-day risk window because hospitalizations were, on average, documented within 2 days of a positive test for the Delta cohort and within 1 day of a positive test for the Emergent Omicron cohort. The 3-day risks of these outcomes were compared between propensity-score matched Emergent Omicron and Delta cohorts. Overall risk, risk ratios, and 95% confidence interval (CI) were calculated. The same 3-day outcomes were compared between the two different Delta cohorts that were propensity-score matched for the same covariates. Separate analyses were performed on patients stratified into five age groups including three pediatric groups (0-4, 5-11, 12-17 years), adults (18-64 years), and older adults (≥ 65 years). All statistical tests were conducted on 12/27/2021 within the TriNetX Analytics Platform with significance set at p-value < 0.05 (two-sided).

## Results

### Patient characteristics

The characteristics of the Emergent Omicron and Delta cohorts before and after propensity-score matching are shown in **Table 1**. Patients from the Emergent Omicron cohort were similar in age to those from the Delta cohort (average age: 36.4 vs 36.1 years), differed in gender and racial and ethnic compositions, had fewer comorbidities and adverse social determinants of health. The usage patterns of many COVID-19 related medications also differed between the two cohorts. Vaccinations documented in patient’s EHRs were lower in the Emergent Omicron cohort than in the Delta cohort, although booster status did not differ. Of patients in the Emergent Omicron vs. Delta cohort, 9.6% vs. 10.4% were 0-4 years old, 9.3% vs. 11.9% were 5-11 years old, 8.8% vs. 8.9% were 12-17 years old, 55.3% vs. 50.6% were 18-64 years old, and 15.8% vs. 17.6% were ≥ 65 years old. After propensity-score matching for variates in the table, the differences between the cohorts decreased or were eliminated.

**Table 1.** Characteristics of the Emergent Omicron cohort and the Delta cohort before and after propensity matching. Emergent Omicron cohort – Patients who contracted SARS-CoV-2 infection between 12/15/2021–12/24/2021 and had no prior SARS-CoV-2 infection. Delta cohort – Patients who contracted SARS-CoV-2 infection between 9/1/2021–11/15/2021 and had no prior SARS-CoV-2 infection. Race and ethnicity as recorded in the TriNetX EHR database and were included in the study because they have been associated with both infection risk and severe outcomes of SARS-CoV-2 infections. P-value – significance between the two cohorts based on two-tailed two-proportion z-test conducted within the TriNetX Network.

|  | Before Matching | After Matching |
| --- | --- | --- |

**Table 1.** Characteristics of the Emergent Omicron cohort and the Delta cohort before and after propensity matching.
|  | **Emergent**<br>**Omicron**<br>**cohort**<br>**(12/15/2021-**<br>**12/24/2021)** | **Delta**<br>**Cohort**<br>**(9/1/2021-**<br>**11/15/2021)** | **P-**<br>**value** | **Emergent**<br>**Omicron**<br>**cohort**<br>**(12/15/2021-**<br>**12/22/2021)** | **Delta**<br>**cohort**<br>**(9/1/2021-**<br>**11/15/2021)** | **P-**<br>**value** |
| --- | --- | --- | --- | --- | --- | --- |
| **Total number of**<br>**patients** | 14,054 | 563,884 |  | 14,040 | 14,040 |  |
| **Age (years, mean±SD)** | 36.4 ± 24.3 | 36.1 ± 25.3 | 0.17 | 36.4 ± 24.3 | 36.1 ± 24.4 | 0.45 |
| **Sex (%)** |  |  |  |  |  |  |
| Female | 55.4 | 53.9 | < .001 | 55.4 | 55.2 | 0.83 |
| Male | 44.6 | 46.0 | < .001 | 44.5 | 44.7 | 0.83 |
| **Ethnicity (%)** |  |  |  |  |  |  |
| Hispanic/Latinx | 4.8 | 8.8 | < .001 | 4.8 | 4.6 | 0.32 |
| Not Hispanic/Latinx | 45.4 | 62.0 | < .001 | 45.4 | 44.8 | 0.30 |
| Unknown | 48.8 | 28.2 | < .001 | 48.8 | 49.6 | 0.19 |
| **Race (%)** |  |  |  |  |  |  |
| African<br>American/Black | 17.1 | 15.0 | < .001 | 17.1 | 16.7 | 0.37 |
| Asian | 1.8 | 2.5 | < .001 | 1.8 | 1.8 | 1.00 |
| White | 57.3 | 61.8 | < .001 | 57.3 | 56.3 | 0.08 |
| Unknown | 22.6 | 19.8 | 0.05 | 22.6 | 23.9 | 0.01 |
| **Adverse Social**<br>**determinants of health**<br>**(%)** | 2.3 | 3.2 | < .001 | 2.3 | 2.0 | 0.19 |
| **Comorbidities (%)** |  |  |  |  |  |  |
| Hypertension | 14.6 | 17.8 | < .001 | 14.6 | 13.7 | 0.02 |
| Heart diseases | 3.7 | 4.7 | < .001 | 3.7 | 3.3 | 0.06 |
| Cerebrovascular<br>diseases | 2.2 | 3.4 | < .001 | 2.2 | 2.1 | 0.59 |
| Obesity | 10.6 | 11.3 | 0.01 | 10.6 | 9.8 | 0.02 |
| Type 2 diabetes | 5.6 | 7.3 | < .001 | 5.6 | 5.3 | 0.29 |
| Cancers | 11.4 | 13.6 | < .001 | 11.4 | 10.3 | 0.002 |
| Chronic respiratory<br>diseases | 10.7 | 13.1 | < .001 | 10.7 | 10.2 | 0.18 |
| Liver diseases | 2.1 | 3.5 | < .001 | 2.1 | 2.0 | 0.67 |
| Chronic kidney disease | 2.2 | 3.2 | < .001 | 2.2 | 2.1 | 0.30 |
| Blood disorders<br>involving immune<br>mechanisms | 10.3 | 12.7 | < .001 | 10.2 | 9.5 | 0.03 |
| HIV infection | 0.16 | 0.23 | 0.08 | 0.16 | 0.16 | 0.88 |
| Dementia | 0.3 | 0.5 | < .001 | 0.3 | 0.3 | 0.91 |
| Substance use disorders | 7.7 | 9.3 | < .001 | 7.7 | 7.5 | 0.54 |
| Depression | 6.6 | 8.6 | < .001 | 6.7 | 6.0 | 0.02 |
| Anxiety | 12.1 | 13.9 | < .001 | 12.1 | 11.5 | 0.13 |
| Smoking | 1.9 | 2.4 | < .001 | 1.9 | 1.4 | 0.004 |
| Alcohol abuse | 0.9 | 1.4 | < .001 | 0.9 | 0.8 | 0.61 |
| **Organ Transplant**<br>**(%)** | 0.4 | 0.5 | 0.02 | 0.4 | 0.3 | 0.47 |
| **COVID-19** |  |  |  |  |  |  |
| **therapeutics (%)** |  |  |  |  |  |  |
| Dexamethasone | 14.7 | 16.8 | < .001 | 14.7 | 13.8 | 0.03 |
| Remdesivir | 0.43 | 0.39 | 0.39 | 0.43 | 0.40 | 0.64 |
| Hydrocortisone | 8.3 | 8.2 | 0.61 | 8.3 | 7.7 | 0.06 |
| Ibuprofen | 27.8 | 24.8 | < .001 | 27.9 | 26.7 | 0.03 |
| Prednisone | 11.7 | 11.1 | 0.02 | 11.7 | 10.8 | 0.02 |
| Methylprednisolone | 12.4 | 12.2 | 0.40 | 12.4 | 10.9 | < .001 |
| Prednisolone | 4.2 | 5.1 | < .001 | 4.2 | 4.0 | 0.51 |
| Naproxen | 6.9 | 6.1 | < .001 | 6.9 | 6.7 | 0.42 |
| Fluoxetine | 2.7 | 2.6 | 0.18 | 2.7 | 2.7 | 0.71 |
| Fluvoxamine | 0.17 | 0.10 | 0.008 | 0.17 | 0.14 | 0.55 |
| Tocilizumab | 0.1 | 0.1 | 0.63 | 0.09 | 0.07 | 0.53 |
| Casirivimab/<br>Imdevimab | 0.1 | 0.6 | < .001 | 0.1 | 0.2 | 0.25 |
| Ritonavir/ Lopinavir | 0.07 | 0.06 | 0.63 | 0.07 | 0.07 | 1.00 |
| **Vaccination in HCOs (%)** |  |  |  |  |  |  |
| Pfizer | 2.4 | 3.1 | < .001 | 2.4 | 2.2 | 0.29 |
| Moderna | 0.3 | 0.4 | 0.02 | 0.3 | 0.2 | 0.36 |
| J& J | 0.07 | 0.05 | 0.22 | 0.07 | 0.07 | 1.00 |
| Booster | 0.8 | 0.8 | 0.89 | 0.8 | 0.6 | 0.07 |

### Lower risks of 3-day acute negative outcomes in the Emergent Omicron cohort than in the matched Delta cohort

The 3-day risks in the Emergent Omicron cohort (n=14,040) were less than half of those in the matched Delta cohort (n=14,040): ED visit: 4.55% vs. 15.22% (RR: 0.30 [0.28-0.33]); hospitalization: 1.75% vs. 3.95% (RR: 0.44 [0.38-0.52]); ICU admission: 0.26% vs. 0.78% (RR: 0.33 [0.23-0.48]); mechanical ventilation: 0.07% vs. 0.43% (RR: 0.16 [0.08-0.32]) (**Figure 1**, top). On the other hand, there were no marked differences in the 3-day risks between the two matched Delta cohorts where patients in the Delta-2 cohort had their first infection between 11/16/2021–11/30/2021(**Figure 1**, bottom), suggesting that the differences in outcomes between the Emergent Omicron cohort and the Delta cohort are largely attributed to differences in the prevalence of the virus variants.

**Figure 1.**
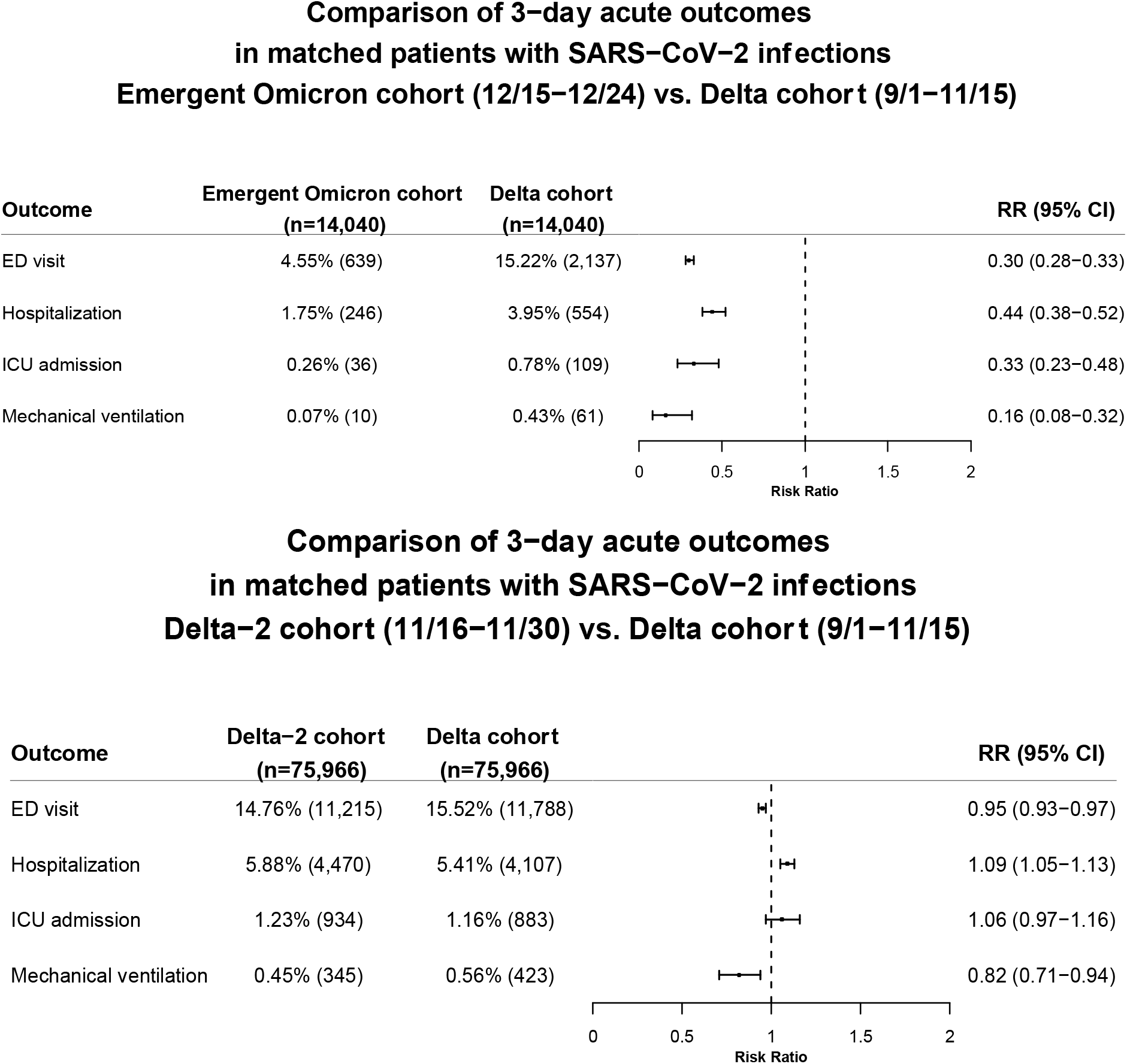
Comparison of 3-day acute outcomes (ED visit, hospitalization, ICU admission, mechanical ventilation) between the matched Emergent Omicron and Delta cohorts (Top panel) and between the two matched Delta cohorts (Bottom panel). Emergent Omicron cohort – Patients who contracted SARS-CoV-2 infection between 12/15/2021–12/24/2021 and had no prior SARS-CoV-2 infection. Delta cohort – Patients who contracted SARS-CoV-2 infection between 9/1/2021–11/15/2021 and had no prior SARS-CoV-2 infection. Delta-2 cohort – Patients who contracted SARS-CoV-2 infection between 11/16/2021–11/30/2021, right before the emergence of the Omicron variant, and had no prior SARS-CoV-2 infection. Cohorts were propensity-score matched for demographics (age, gender, race/ethnicity), socioeconomic factors, transplant procedures, COVID-19 related comorbidities, COVID-19 related medications, and EHR-documented vaccination status.

### Lower risk for 3-day hospitalization and ED admissions in the Emergent Omicron vs matched Delta cohort in children and adults

In children from the Emergent Omicron cohort under 5 years old (n=1,361), the 3-day risk for ED visit was 3.89%, significantly lower than 21.01% in the matched Delta cohort (RR: 0.19 [0.14-0.25]). The 3-day risk for hospitalization was 0.96%, significantly lower than 2.65% in the matched Delta cohort (RR: 0.36 [0.19-0.68]). Lower risks for ED visit and hospitalization were observed for the two other pediatric groups (aged 5-11 years, and 12-17 years), though differences were not significant for hospitalization, which may be due to the small number of 5– 11-year-old and 12–17-year-old who were hospitalized. The Emergent Omicron cohort had less severe disease than the matched Delta cohort for both adults (18-64 years) and older adults (≥ 65 years) (**Figure 2**, top). In contrast, no marked differences were observed between the two matched Delta cohorts for all age groups (**Figure 2**, bottom).

**Figure 2.**
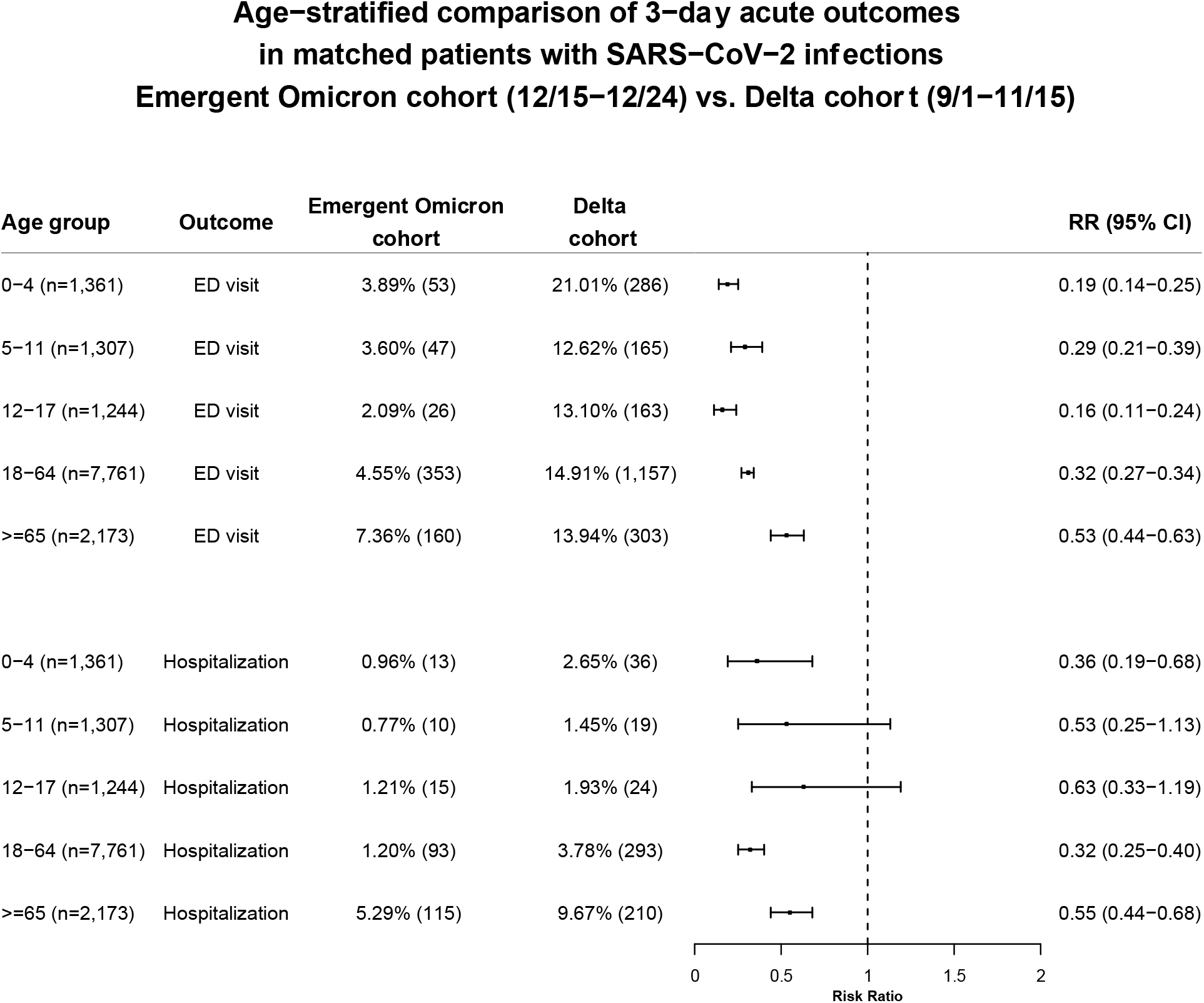
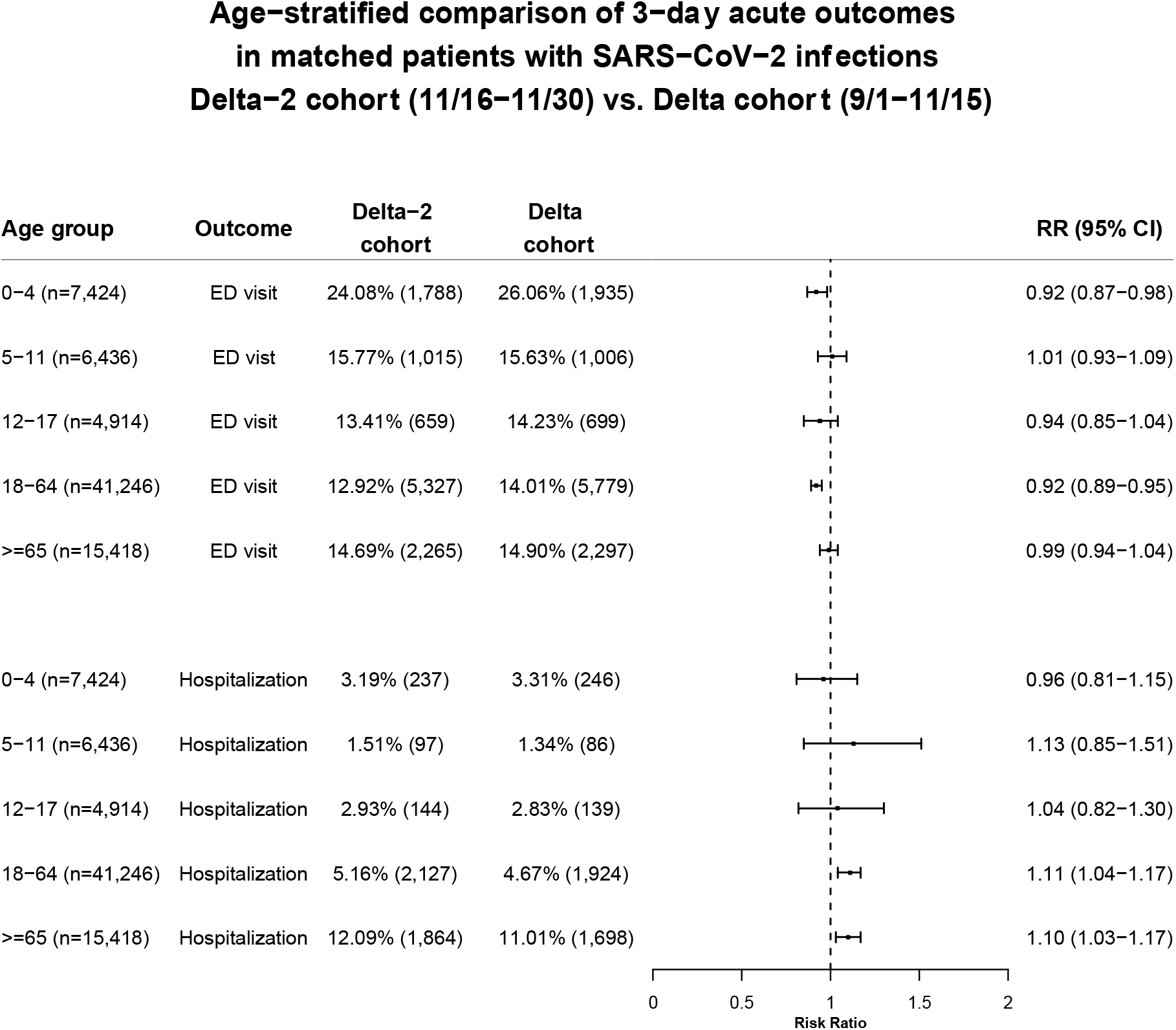
Comparison of 3-day ED visit and hospitalization in children, adults, and older adults between propensity-score matched Emergent Omicron and Delta cohorts (Top panel) and between two matched Delta cohorts (Bottom panel). Emergent Omicron cohort - Patients who contracted SARS-CoV-2 infection between 12/15/2021–12/24/2021 and had no prior SARS-CoV-2 infection. Delta cohort–Patients who contracted SARS-CoV-2 infection between 9/1/2021–11/15/2021 and had no prior SARS-CoV-2 infection. Delta-2 cohort–Patients who contracted SARS-CoV-2 infection between 11/16/2021–11/30/2021, immediately prior to the emergence of the Omicron variant, and had no prior SARS-CoV-2 infection. Cohorts were propensity-score matched for demographics (age, gender, race/ethnicity), socioeconomic factors, transplant procedures, COVID-19 relevant comorbidities and medications, and EHR-documented vaccination status.

## Discussion

In this study using a nation-wide database of EHRs in the US, SARS-CoV-2 infected patients in the period when the Omicron variant emerged were demographically different from and had fewer adverse health conditions than those infected during the previous period when the Delta variant predominated. After matching for age, gender, race/ethnicity, adverse socio-economic determinants of health, COVID-related comorbidities and medications, and EHR-documented vaccination status, the 3-day risk of hospitalization from SARS-CoV-2 infections that occurred during the period when the Omicron variant emerged was 1.75%, less than half of the 3.95% observed for the period when the Delta variant predominated and little or no Omicron variant was recorded. This result is consistent with findings in South Africa^4^, Scotland^5^ and England^6^. In contrast, no major changes in hospitalization were observed for SARS-CoV-2 infections occurring during the two-week Delta variant period (11/16/2021–11/30/2021) that immediately preceded the emergence of the Omicron variant and the 10-week Delta variant period before it (9/1/2021–11/15/2021) indicative of relatively stable outcomes.

This study is among the first to provide evidence of reduced severity from infections that occurred in the period after the emergence of the Omicron variant in the US as compared to the period that preceded it. The 3-day risks from the Emergent Omicron cohort for ED visit was less than one-third of that for the Delta variant (4.55% vs 15.22%); for ICU admission, it was one-third of that for the Delta variant (0.26% vs 0.78%); for mechanical ventilation, it was one-sixth of that for the Delta variant (0.07% vs 0.43%). To test whether these differences in outcomes might represent merely the gradual accrual of immunized members of the population plus increasing availability of treatments designed to prevent hospitalization, we compared outcomes in patients who were first infected between 9/1/2021–11/15/2021 (Delta period) to those in patients who were first infected between 11/16/2021–11/30/2021 (Delta period immediately before Omicron emergence, which we called Delta-2). No marked differences in outcomes were found between these two Delta cohorts after propensity-score matching. These results further suggest that the Omicron variant is likely to be associated with milder disease outcomes than the Delta variant. Mortality risks were not reported in this study because so few deaths occurred within 3 days of infection in both cohorts: 23 (0.16%) vs. 30 (0.21%) among 14,040 patients in the matched Emergent Omicron and Delta cohorts. Longer follow-up times are needed to accurately estimate mortality risk.

Pediatric SARS-CoV-2 infections and hospitalizations are rising in South Africa^11^ and in the US. This is especially concerning given that in the US, children under the age of 5 are not eligible for COVID-19 vaccination, and children ages 5 through 11 are not eligible to receive boosters. This study shows that the risk for hospitalization in unvaccinated children under 5 years old that occurred after the emergence of the Omicron variant was one-third of that during the Delta variant period (0.96% vs. 2.65%) whereas the risk for ED visit was less than one-fifth (3.89% vs. 21.01%), both differences were significant. The same trends were observed for children 5-11 and 12-17 years old. These results suggest that while pediatric SARS-CoV-2 infections and hospitalizations are reportedly rising, the outcomes are milder after the emergence of the Omicron variant as compared to the predominant Delta variant period that preceded it. No significant differences in pediatric outcomes were observed between the two different Delta cohorts, suggesting that the differences in outcomes between the Emergent Omicron cohort and the Delta cohort are largely attributed to differences in the prevalence of the virus variants. The risks for hospitalization and ED visit were higher for children under 5 years old than for children 5-11 years old (0.96% vs 0.77% for hospitalization and 3.89% vs. 3.60% for ED visit), but these differences were not significant. ICU admissions and mechanical ventilation were not examined in the age-stratified groups due to limited sample sizes.

In summary, compared to patients first infected during 9/1-11/15, there is a marked decrease in ED visits, hospitalization and ICU utilization and mechanical ventilation for patients first infected during 12/15/-12/24. No such decreases were observed for patients first infected during 11/16/-11/30, immediately prior to the emergence of the Omicron variant, so there is no evidence of gradual waning of the Delta variant virulence or rapid ramp up of early COVID treatments, which are still not widely available. Though the decrease in disease severity in the Emergent Omicron cohort could reflect increases in vaccination and boosters in the population between November and December, the proportion would have had to be quite substantial to account for the large reductions that we observed. In addition, we did not observe similar decreases in disease severity for infections occurring right before the emergence of the Omicron variant, as might occur with a gradual ramp up of vaccination. Moreover, since we selected persons who had not had prior SARS-CoV2, it is unlikely that the milder results occurred because of persistence of viral mRNA from prior infection and not from new, acute infection. Taken together, it is likely that the emergence and increased prevalence of Omicron entrains milder diseases and is a major contributor to the observed decreases in disease severity. The estimated prevalence of the Omicron variant during 12/15-12/24 was only 22.5-58.6%, suggesting that the outcomes for the Omicron variant may be found to be even milder than what we report here as the prevalence of the Omicron variant increases. Findings from this study, combined with the approval of COVID-19 medications and the increases in numbers of people who are vaccinated and boosted, present a more encouraging picture of the population response to COVID infection.

Our study has several limitations: First, the observational, retrospective nature of this study of patient EHR data could introduce selection, information, testing, reporting and follow up issues. However, because we compared the different population all from the TriNetX dataset, these issues should not significantly affect the relative risk analyses. Second, patients in the TriNetX EHR database are those who had medical encounters with healthcare systems contributing to the TriNetX Platform and do not necessarily represent the entire US population. Therefore, results from the TriNetX platform need to be validated in other populations. Third, both the Emergent Omicron and Delta cohorts in our study were defined based on CDC’s national genomic sequence surveillance. The Emergent Omicron cohort likely contained some infections with the Delta variant, but this admixture would tend to reduce the observed differences. However, our findings of reduced hospitalization in the Emergent Omicron cohort compared to the Delta cohort is consistent with findings from Africa^4^, Scotland^5^, and England^6^ that were based on genomic sequences, and goes further to indicate that the severity in hospitalization is reduced. The fact that there were no marked differences between the two Delta cohorts yet significant differences between the Emergent Omicron and Delta cohorts further corroborates that the differences in outcomes for infections occurring between 12/15/2021–12/24/2021 were likely to be caused largely by the emerging Omicron variant. Fourth, we are unable to determine with certainty the vaccination status of the infected individuals as the SARS-CoV2 vaccination information in TriNetX is incomplete. The vaccination rate documented in TriNetX is only about 2%, whereas the reported vaccination rates during this time period^12^ indicate that most patients in the study population were likely to have been vaccinated. However, the cohorts were propensity-score matched for EHR-documented vaccination status including vaccine types and booster along with demographics, adverse socioeconomic determinants of health and comorbidities, some of which are associated with vaccination acceptance^13,14^. Finally, the findings apply only to infections that occurred in the US between 12/15/2021– 12/24/2021. Given the fast increases in the prevalence of the Omicron variant, including the potential for further viral mutations, future studies are imperative to closely follow up disease severity as well as longer term outcomes associated with infections from Omicron or other variants.

In summary, our analysis indicates that the emergence of the Omicron variant in the US was associated with significantly milder disease in the early phase of infection as compared to the Delta variant period that preceded it. Despite this encouraging result, further studies are needed to monitor the longer-term acute consequences from Omicron infection, the propensity for development of “long COVID”, the rapidity of spread, potential for mutation, and how the vaccines, boosters, or prior infections alter clinical responses. Additionally, although infections from the Omicron variant, based on this analysis, appear to be milder, because of Omicron’s increased transmissibility, the overall number of ED visits, hospitalizations, ICU admissions, and mechanical ventilator patients may still be greater with the Omicron variant then the Delta variant.

## Data Availability

All data produced in the present work are contained in the manuscript

## Contributors

RX conceived, designed, and supervised the study, performed literature review and reviewed and drafted the manuscript. LW conducted all the experiments, performed data analysis and prepared tables and figures. NAB, PBD, DCK, and NDV critically contributed to study design, result interpretation and manuscript preparation. We confirm the originality of content.

## Acknowledgments

We acknowledge support from National Institute on Aging (grants nos. AG057557, AG061388, AG062272), National Institute on Alcohol Abuse and Alcoholism (grant no. R01AA029831), National Institute on Drug Abuse (grant no. UG1DA049435), the Clinical and Translational Science Collaborative (CTSC) of Cleveland (grant no. 1UL1TR002548-01), National Cancer Institute Case Comprehensive Cancer Center (R25CA221718, P30 CA043703, P20 CA2332216).

## Meeting Presentation

No

## References

1. CDC. COVID Data Tracker [Internet]. [cited 2021 Dec 23];Available from: https://covid.cdc.gov/covid-data-tracker/#variant-proportions

2. CDC. Omicron Variant: What You Need to Know [Internet]. 2021.[cited 2021 Dec 24];Available from: https://www.cdc.gov/coronavirus/2019-ncov/variants/omicron-variant.html

3. Ledford H. How severe are Omicron infections? Nature 2021;600(7890):577–8.

4. Wolter N, Jassat W, Walaza S, et al. Early assessment of the clinical severity of the SARS-CoV-2 Omicron variant in South Africa [Internet]. bioRxiv. 2021;Available from: http://medrxiv.org/lookup/doi/10.1101/2021.12.21.21268116

5. Sheikh A, Kerr S, Woolhouse M, McMenamin J, Robertson C. Severity of Omicron variant of concern and vaccine effectiveness against symptomatic disease: national cohort with nested test negative design study in Scotland. 2021 [cited 2021 Dec 23];Available from: https://www.pure.ed.ac.uk/ws/files/245818096/Severity_of_Omicron_variant_of_concern_and_vaccine_effectiveness_against_symptomatic_disease.pdf

6. Report 50 - Hospitalisation risk for Omicron cases in England [Internet]. [cited 2021 Dec 23];Available from: https://www.imperial.ac.uk/mrc-global-infectious-disease-analysis/covid-19/report-50-severity-omicron/

7. TriNetX [Internet]. 2021.[cited 2021 Dec 17];Available from: https://trinetx.com/

8. COVID-19 and People at Increased Risk [Internet]. 2021 [cited 2021 Dec 17];Available from: https://www.cdc.gov/drugoverdose/resources/covid-drugs-QA.html

9. CDC. People with Certain Medical Conditions [Internet]. 2021 [cited 2021 Dec 17];Available from: https://www.cdc.gov/coronavirus/2019-ncov/need-extra-precautions/people-with-medical-conditions.html

10. Hospitalized Adults: Therapeutic Management [Internet]. [cited 2021 Dec 24];Available from: https://www.covid19treatmentguidelines.nih.gov/management/clinical-management/hospitalized-adults--therapeutic-management/

11. Cloete J, Kruger A, Masha M, et al. Rapid rise in paediatric COVID-19 hospitalisations during the early stages of the Omicron wave, Tshwane District, South Africa [Internet]. bioRxiv. 2021;Available from: http://medrxiv.org/lookup/doi/10.1101/2021.12.21.21268108

12. Ritchie H, Mathieu E, Rodés-Guirao L, et al. Coronavirus Pandemic (COVID-19). Our World in Data [Internet] 2020 [cited 2021 Dec 27];Available from: https://ourworldindata.org/coronavirus

13. Soares P, Rocha JV, Moniz M, et al. Factors Associated with COVID-19 Vaccine Hesitancy. Vaccines (Basel) [Internet] 2021;9(3). Available from: http://dx.doi.org/10.3390/vaccines9030300

14. Tsai R, Hervey J, Hoffman K, et al. COVID-19 vaccine hesitancy and acceptance among individuals with cancer, autoimmune diseases, and other serious comorbid conditions: A cross-sectional internet-based survey. JMIR Public Health Surveill [Internet] 2021;Available from: http://dx.doi.org/10.2196/29872

